# A statistical framework to identify gene-gene interactions underlying multiple dichotomous phenotypes from genotype data

**DOI:** 10.1101/2025.11.09.25339828

**Authors:** Siru Wang, Xuhui Zhu, Yu Li, Gengjie Jia

**Author notes:** Equal contributing first authors (Siru Wang, Xuhui Zhu).

## Abstract

Identifying gene-gene (G × G) interactions across multiple dichotomous phenotypes is challenging due to the extreme sparsity of SNP-derived interaction matrices and reduced statistical power by binary outcomes. Existing G×G association methods are restricted to either single or multiple continuous phenotypes. Here we introduce GiMat (Gene Interaction and Multiple-phenotype Association Test), a statistical framework that extends multivariate kernel regression to model G × G interactions jointly across dichotomous phenotypes, while explicitly capturing homogeneous and heterogeneous interaction effects. Extensive simulations demonstrate that GiMat controls type I error conservatively and adapts power flexibly to different types of relationships between interaction effects and phenotypes. Applied to type 2 diabetes and hypertension comorbidity in the UK Biobank, GiMat identified four previously unreported G×G interaction pairs associated with both phenotypes. This scalable framework enables robust discovery of complex genetic interactions underlying multiple correlated phenotypes.

## Introduction

Understanding how genes jointly influence disease risk is essential for deciphering complex phenotypes and guiding precision interventions. Genome-wide association studies (GWAS) have uncovered numerous genetic variants linked to complex phenotypes^1,2^, yet traditional single-phenotype analyses explain only a fraction of disease heritability. Gene-gene (G×G) interactions, in which the effect of one gene modifies another^3^, are thought to play a crucial role in disease pathogenesis and may account for some of this “missing heritability”^4^. However, detecting G×G interactions across multiple dichotomous phenotypes remains particularly challenging^5,6^ due to the extreme sparsity of SNP-derived interaction matrices and reduced statistical power by binary outcomes.

Existing methods for testing G×G interactions fall broadly into non-parametric and parametric classes. Non-parametric methods such as multifactor dimension reduction (MDR)^7,8^ can detect higher-order interactions in dichotomous phenotypes but lack interpretability. In contrast, parametric methods like generalized linear models provide interpretable effect estimates but struggle with higher-order or sparse interactions^9,10^. Most methods were developed for single or continuous phenotypes, not for multi-phenotype binary outcomes, where sparsity and heterogeneity issues are amplified.

The emergence of multi-locus, multi-phenotype analyses reflect recognition that correlated phenotypes share intertwined molecular mechanisms^11,12^. Dual-kernel approaches such as GAMuT^13^, DKAT^14^, and score-based MSKAT^15^ test cross-phenotype effects but are restricted to continuous outcomes and rely on assumptions (*e*.*g*., homogeneous effects) that, when violated, reduce power^16^. In addition, DKAT and MSKAT incur heavy computational burdens at large sample sizes, with DKAT performing best when phenotype dimensionality exceeds sample size.

Here we introduce the Gene Interaction and Multiple-phenotype Association Test (GiMat), a statistical framework specifically designed to uncover G×G interactions across multiple dichotomous phenotypes. GiMat extends multivariate regression by incorporating flexible phenotype kernels to model homogeneous or heterogeneous interaction effects and applies a saddlepoint approximation^17-20^ to calibrate score statistics under imbalanced case-control ratios. In extensive simulations and comorbidity applications, GiMat demonstrated conservative type I error control and capability of discovering previously unreported G×G interactions.

## Results

### Type I error and statistical power across varied phenotype prevalence, correlation, and genetic effect scenarios

We conducted a series of simulations to evaluate GiMat performance under two genetic-effect settings: one without main genetic effects (*α*_1_ = *α*_2_ = 0; see Simulation design in Methods) and one with main genetic effects (*α*_1_ ≠ 0, *α*_2_ ≠ 0). In each scenario, two dichotomous phenotypes were generated at correlation 0.5 with prevalences of 5%, 10%, 20%, and 50%. Empirical type I errors—calculated as the proportion of *p*-values below 0.05, 0.01, 0.005, 0.001, and 0.0005 (Supplementary Table 1)—were consistently conservative across all settings. The *minP* kernel yielded the lowest type I error rates, while other kernels showed comparable performance. Lower disease prevalence further reduced type I error. With main effects present, type I errors became markedly deflated, reaching zero at the 0.0005 threshold across all prevalence levels, reflecting the dominance of non-interaction effects on phenotype generation.

To test sensitivity to phenotype correlation, we repeated simulations with correlations of 0.3 and 0.75 under 20% prevalence (Supplementary Table 2). Type I errors increased slightly at lower correlation, consistent with weaker redundancy between phenotypes. High correlation makes the phenotypes more similar in their associations with G× G interactions, dampening false positives, whereas weaker correlation leads to more independent noise and slightly increased type I error.

Power was then compared across different kernels under both genetic-effect scenarios with phenotype correlation fixed at 0.5. We simulated 20%, 60%, and 100% of G×G interactions as causal, with either all-positive G×G interaction effects or an equal mix of half-positive and half-negative G×G interaction effects (Fig. 1). Statistical power was defined as the proportion of *p*-values below the nominal significance level (*θ* = 0.05). Without main genetic effects, the *Hom* kernel showed modestly higher power for all-positive effects of G × G interactions compared to the power of other kernels. However, the gains in power of *Hom* kernel decreased as the proportion of causal interactions increased (10/0/0). Under mixed-direction effects, the power of *Hom* kernel decreased noticeably and the power of *Het* kernel increased significantly; while the *PhC, GAM*, and *minP* kernels remained intermediate and stable. With main genetic effects present, overall power decreased. Under the all-positive interaction setting (interaction effects were homogeneous across phenotypes), *Hom* kernel produced significant gains in power compared to *minP, PhC*, and *Het*; in contrast, with mixed-direction effects (half-positive and half-negative), *Het* achieved the highest power while *Hom* demonstrated the lowest, with differences most marked at low interaction proportions.

**Fig. 1.**
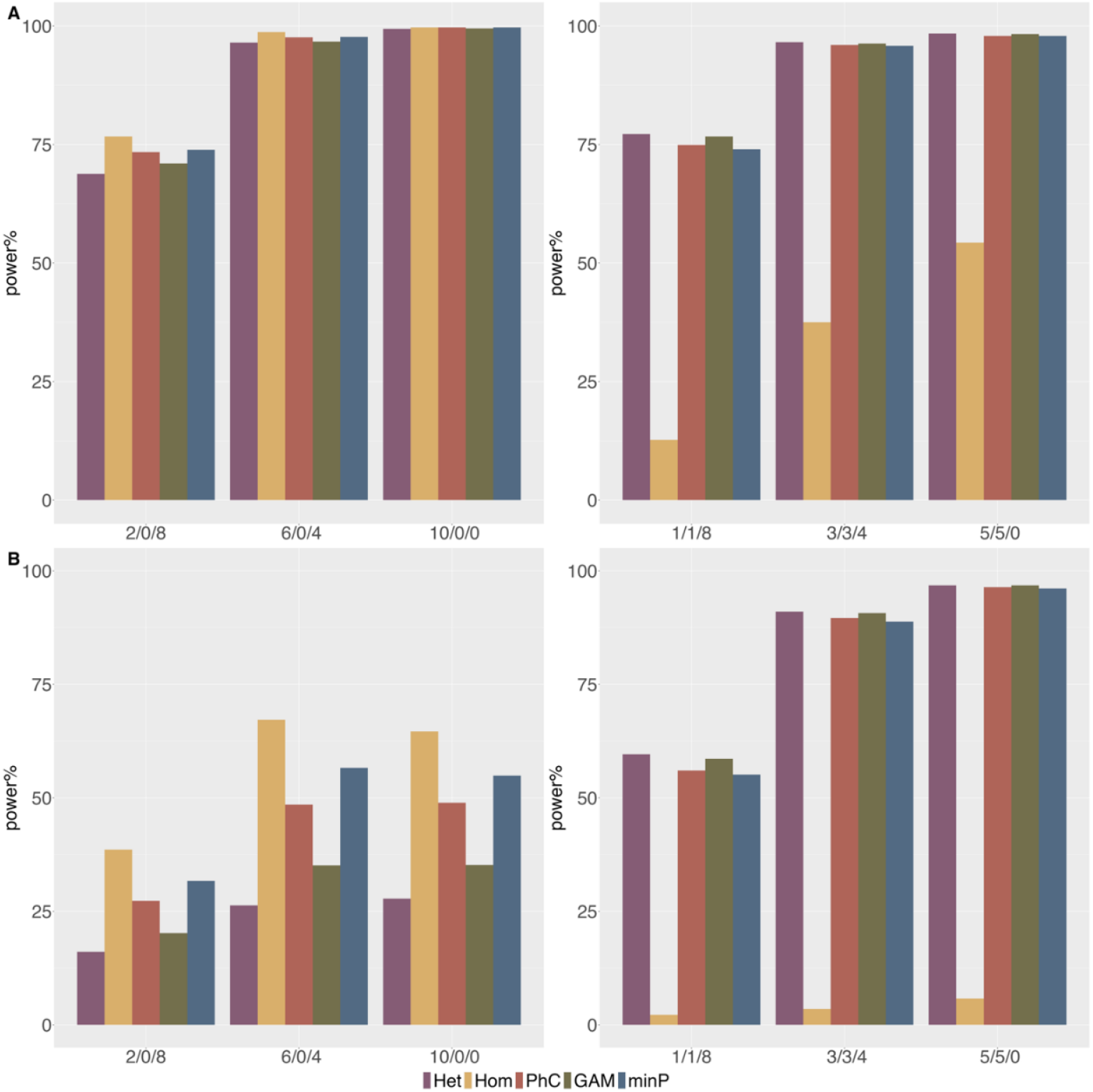
Empirical power of *Het, Hom*, and *PhC* kernels at 20% disease prevalence. Bars depict empirical power, defined as the proportion of simulations with *p*-values < 0.05, for different configurations of interaction effect directions. The x-axis (format: “+/-/0”) indicates the number of interaction variants with positive, negative, and null effects, respectively. Panel A: scenario without main genetic effects; Panel B: scenario with both main genetic effects and causal interactions effects acting on different phenotypes.

Finally, we examined power as a function of interaction-effect magnitude by varying causal G×G effects from 0 to 0.4 in steps of 0.05 under the scenario where different G×G effects influenced different phenotypes (Supplementary Fig. 1). With all-positive effects and main genetic effects, power increased with effect size across all kernels but stayed below 75%, with *Hom* consistently highest due to aligned interaction directions across phenotypes. In contrast, with mixed-direction effects, power gains were greater for *Het, PhC*, and *GAM* as effect size increased, whereas *Hom* improved only modestly, underscoring its sensitivity to effect concordance.

### Application to detecting G×G interactions in T2D-HTN comorbidity

To investigate G×G interactions underlying the comorbidity of T2D and HTN, we curated independent genome-wide significant SNPs (*p*-value < 5×10^−8^) and their mapped genes for T2D and HTN from published GWAS. Each T2D risk gene was paired with each HTN risk gene, generating all plausible cross-disease combinations for interaction testing. Because onset timing may mark mechanistic heterogeneity, UK Biobank participants were stratified into four subgroups based on the difference in diagnosis ages for T2D versus HTN: (−∞, −5], (−5, 0], (0, 5], and (5, +∞) years (Supplementary Table 3). GiMat was then applied separately to each subgroup using the *Hom, Het, PhC, GAM*, and *minP* kernels, yielding 20 sets of test statistics.

Across these analyses GiMat identified four G × G pairs that were both statistically significant and supported by independent molecular evidence in the IntAct^21^ and STRING^22^ databases (Supplementary Table 4), with the complete set of results provided in Supplementary Data 1. HLA-DQA1 and HLA-DQB1 encode the *α*- and β-chains of the MHC-II complex forming HLA-DQ8^23^, a well-established type I diabetes risk factor^23^ confirmed structurally in the human insulin peptide-HLA-DQ8 complex, reinforcing the biological plausibility of this interaction. After accounting for multiple testing using Bonferroni correction (*p*-value < 7.35×10^−4^), each age-difference subgroup yielded at least one significant interaction (Table 1): SLC30A8-MTHFR (most significant under the *Hom* kernel, followed by *Het*), INS-RP1L1 (significant under *Het, Hom*, and *GAM*), EHMT2-HLA-DQB1 (significant under all kernels except *PhC*), and DXO-ZBTB10 (detected by *PhC* and *Het*). Three of the four pairs involved one gene tagged by a single SNP paired with a multi-SNP gene, reflecting the heterogeneous architecture of cross-locus interactions.

**Table 1.**
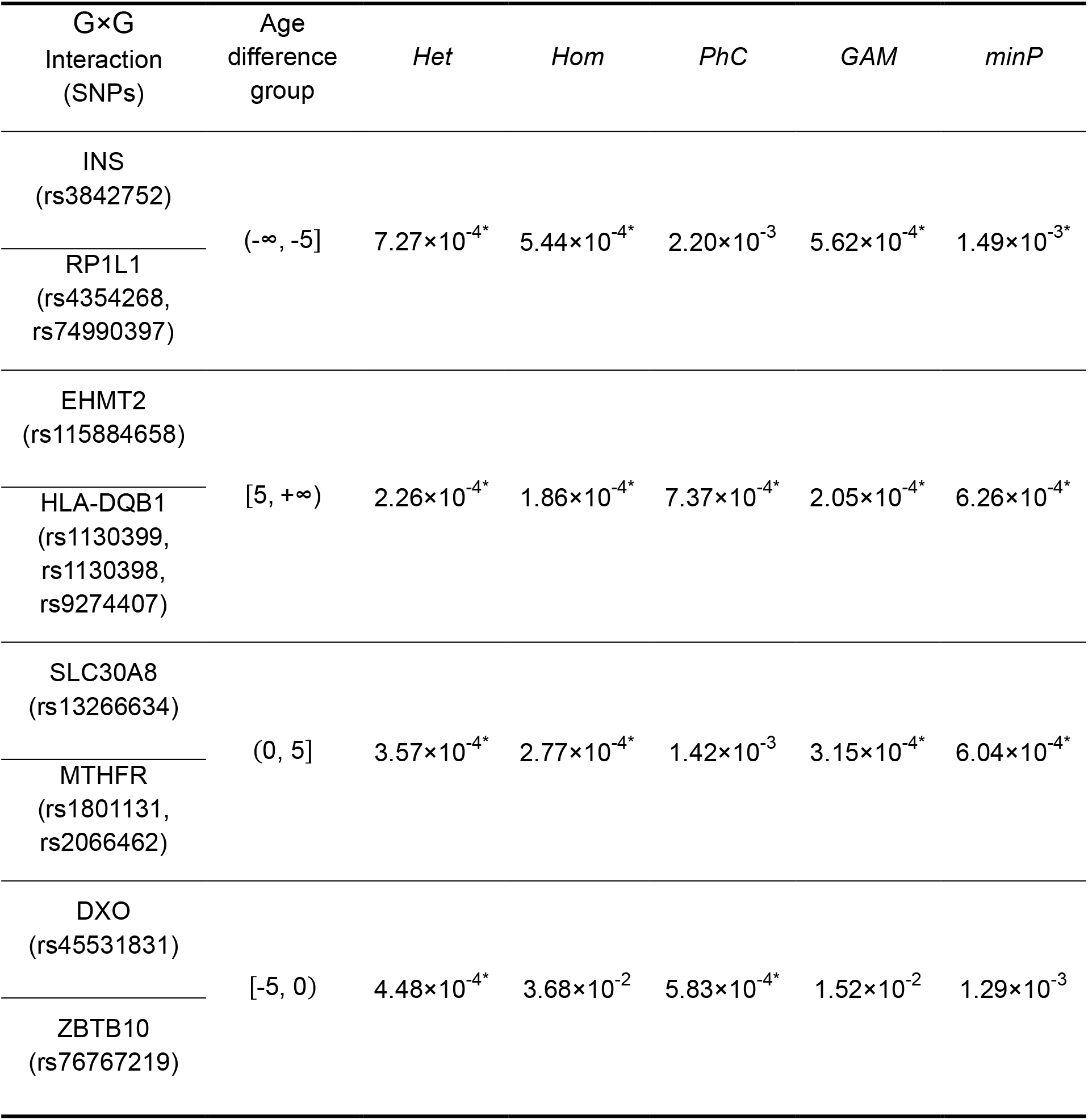
Significant and suggestive G×G interactions associated with T2D and HTN. This table reports G × G interactions with *p*-values below the Bonferroni-corrected threshold (*p*-value < 7.35 × 10^−4^) in any of the GiMat kernel tests: *Het* (heterogeneous kernel), *Hom* (homogeneous kernel), *PhC* (phenotype covariance kernel), *GAM* (gene association with multi-trait test), and *minP* (minimum *p*-value omnibus test). An asterisk (*) indicates *p*-values surpassing the Bonferroni significance threshold. Each row corresponds to a G×G interaction, with the first gene (associated with T2D) and the second gene (associated with HTN) reported under “G×G interaction (SNPs)” alongside genotyped SNP identifiers in parentheses. The “Age difference group” column specifies the participant subgroup in which the significant association was observed, defined by the interval of T2D onset age minus HTN onset age.

| G×G<br>Interaction<br>(SNPs) | Age<br>difference<br>group | *Het* | *Hom* | *PhC* | *GAM* | *minP* |
| --- | --- | --- | --- | --- | --- | --- |
| INS<br>(rs3842752) | (-∞, -5] | $7.27 \times 10^{-4*}$ | $5.44 \times 10^{-4*}$ | $2.20 \times 10^{-3}$ | $5.62 \times 10^{-4*}$ | $1.49 \times 10^{-3*}$ |
| RP1L1<br>(rs4354268,<br>rs74990397) |  |  |  |  |  |  |
| EHMT2<br>(rs115884658) | [5, +∞) | $2.26 \times 10^{-4*}$ | $1.86 \times 10^{-4*}$ | $7.37 \times 10^{-4*}$ | $2.05 \times 10^{-4*}$ | $6.26 \times 10^{-4*}$ |
| HLA-DQB1<br>(rs1130399,<br>rs1130398,<br>rs9274407) |  |  |  |  |  |  |
| SLC30A8<br>(rs13266634) | (0, 5] | $3.57 \times 10^{-4*}$ | $2.77 \times 10^{-4*}$ | $1.42 \times 10^{-3}$ | $3.15 \times 10^{-4*}$ | $6.04 \times 10^{-4*}$ |
| MTHFR<br>(rs1801131,<br>rs2066462) |  |  |  |  |  |  |
| DXO<br>(rs45531831) | [-5, 0) | $4.48 \times 10^{-4*}$ | $3.68 \times 10^{-2}$ | $5.83 \times 10^{-4*}$ | $1.52 \times 10^{-2}$ | $1.29 \times 10^{-3}$ |
| ZBTB10<br>(rs76767219) |  |  |  |  |  |  |

We next compared the distributions of significant *p*-values (*p*-value < 0.05) across age-difference subgroups using Wilcoxon signed-rank tests (Supplementary Table 5). The (0, 5] subgroup consistently produced stronger interaction signals than other age subgroups, supporting the hypothesis that a shorter interval between T2D and HTN onset reflects and perhaps was attributable to stronger underlying genetic interactions.

Functional coherence of the four significant pairs was evaluated using RNA-seq data from 54 non-diseased tissues of nearly 1,000 individuals in the Adult Genotype-Tissue Expression (GTEx) project^22,23^. Pearson correlations and FDR-adjusted *p*-values were computed for each gene pair per tissue (Supplementary Fig. 2). RP1L1 and INS showed specific co-expression in adrenal gland, whereas the other pairs displayed broader but less tissue-specific expression, across more than 10 tissue types. Literature and database mining reinforced biological plausibility: DXO is linked to melanoma via fastBAT^24^; ZBTB10 is nearest to a keratinocyte carcinoma risk locus (*rs*10102591)^25^; SLC30A8 encodes the zinc transporter ZnT8, essential for *β*-cell zinc storage and Ca^2+^ channel activity^26^; and MTHFR, a folate-cycle enzyme for which zinc is a cofactor^27^, was detected as interacting with SLC30A8. Although no direct SLC30A8-MTHFR interaction has been reported, HumanNet analysis^28^ revealed both genes connect through SYT2, suggesting an indirect protein link^28-30^. Likewise, EHMT2 and HLA-DQB1 were found among the shared susceptibility genes for T2D and inflammatory bowel disease^31^; HumanNet identified GATA3 as a bridging factor, with EHMT2 interacting with GATA3 at the protein level^28,32^ and GATA3 co-functionally linked to HLA-DQB1, indicating a potential EHMT2-GATA3-HLA-DQB1 regulatory axis.

Finally, we applied the Gene-Gene Interaction Predictor Neural Network (GGIPNN)^33^ to test functional relatedness. Using 200-dimensional gene embeddings derived from the Gene Expression Omnibus (GEO) database^34^ and 30 independent runs, GGIPNN consistently predicted INS-RP1L1 as interacting (30/30 runs), SLC30A8-MTHFR and DXO-ZBTB10 in the majority (19/30) and EHMT2-HLA-DQB1 in 13/30 runs. This convergence of statistical, expression, and network evidence underscores the biological relevance of the interaction signals uncovered by GiMat.

## Discussion

Comorbidity often reflects shared or interacting genetic etiology. Here we introduced GiMat, a statistical framework for testing G×G interactions across multiple dichotomous phenotypes. GiMat extends the Multi-SKAT architecture^35^ to flexibly model heterogenous and homogeneous interaction effects using multiple kernel structures. When interaction effects on phenotypes are assumed independent, the heterogeneous (*Het*) kernel maximizes sensitivity; when effects are assumed directionally aligned, the homogeneous (*Hom*) kernel is optimal. For scenarios lacking prior biological information to guide kernel choice, GiMat incorporates an extended Copula-based *p*-value combination^35^ to aggregate results across kernels, thereby enabling robust inference without sacrificing power.

Extensive simulations showed that GiMat conservatively controls type I error across a broad range of scenarios. When phenotypes are unrelated to the same genes or linked to distinct genes, type I error rates remained deflated across kernels, even at stringent thresholds. Regardless of whether main genetic effects present, power analyses showed that kernel choice influences detection sensitivity: when G×G effects on phenotypes are independent (half-positive, half-negative), the *Hom* kernel yielded the lowest power, whereas under homogeneous effects with shared genetic influences on both phenotypes, *Hom* produced the highest power compared to other kernels, particularly at high proportions of causal interactions.

Applied to UK Biobank data, GiMat uncovered four previously unreported G×G interactions between T2D and HTN risk genes (SLC30A8–MTHFR, DXO– ZBTB10, INS–RP1L1, and EHMT2–HLA-DQB1), stratified by onset-age intervals. This illustrates GiMat’s capacity to capture temporally stratified genetic effects underlying comorbidity. The SLC30A8-MTHFR interaction, for example, implicates potential crosstalk between zinc transport and folate metabolism, and interaction effects were 2.3-fold stronger when T2D onset preceded HTN by less than five years, suggesting that early metabolic dysregulation in T2D may prime hypertensive pathophysiology.

Despite these advances, several practical challenges remain. First, GiMat currently targets common variants (MAF > 1%), limiting sensitivity to rare variant interactions. Incorporating STAAR-like or MultiSTAAR mixed-model frameworks could extend rare variant detection^36^ and accommodate whole-genome sequencing data and complex population structures^37^. Second, sample-size requirements also grow rapidly with phenotype dimensionality; in our UK Biobank analysis, stringent case-control matching retained only 4.8% of eligible individuals, underscoring the need for adaptive sampling strategies such as POLMM-GENE^38^. Finally, the combinatorial explosion of gene-pair tests—even after restricting to known risk loci (1,940 pairs)—necessitates intelligent filtering. Integrating machine-learning-based pre-screening or tissue-specific interaction kernels derived from GTEx eQTL data^23^ could prioritize biologically relevant pairs and enable pathway-focused comorbidity modelling.

Statistical interactions alone do not guarantee biological relevance. Some, such as DXO–ZBTB10, may reflect regulatory cascades rather than direct molecular contact, underscoring the need for multi-tiered validation. Critical next steps include replication in independent cohorts, perturbation-based functional assays (*e*.*g*., CRISPR, qPCR), and structural modeling with AlphaFold-Multimer^39^ to assess binding potential (*e*.*g*., pDockQ scores^40^). Among the four GiMat-detected pairs, DXO–ZBTB10 and EHMT2–HLA-DQB1 exhibited the highest predicted binding confidences (pDockQ = 0.742 and 0.741), whereas INS–RP1L1, although statistically significant (*Het p*-value = 3.57 × 10^−4^), showed a lower pDockQ (0.573), suggesting indirect or context-dependent coupling. These findings highlight the value of integrating pathway enrichment (*e*.*g*., KEGG^41^, Reactome^42^), tissue co-expression (*e*.*g*., GTEx^23^), and epigenomic overlap (*e*.*g*., ChIP-seq^43^) to distinguish causal interactions from correlative signals.

In summary, GiMat provides a flexible and scalable statistical framework for uncovering G×G interactions across multiple dichotomous phenotypes using SNP-derived genotype data. By combining kernel-based multi-phenotype regression with saddlepoint-calibrated score tests, GiMat robustly controls type I error, adapts to heterogeneous or homogeneous interaction patterns, and recovers biologically plausible signals. Although developed for common variants, its architecture can be extended to rare variants and other interaction types such as gene-environment or environment-environment interactions. This scalability and interpretability fill a critical methodological gap, enabling deeper dissection of the genetic architecture of comorbidities and informing precision medicine.

## Online Methods

### Notation

We developed a statistical approach, the Gene Interaction and Multiple-phenotype Association Test (GiMat), especially designed for testing associations between gene-gene (G×G) interactions and multiple dichotomous phenotypes. Suppose data are available from *n* unrelated individuals genotyped across a genomic region containing *p* common variants (minor allele frequency > 5%) and measured on *K* dichotomous phenotypes. Let *Y* = (***y***_**1**_, ***y***_**2**_, …, ***y***_***K***_), where ***y***_***k***_ = (*y*_1*k*_, *y*_2*k*_, …, *y*_*nk*_)^*T*^ denotes the *n* × 1 vector of the *k*-th dichotomous phenotypes across *n* individuals. Let *X* be the *n* × *m* matrix of *m* covariates, including age, sex, and genetic principal components to account for population structure. For clarity, we focus on two gene regions genotyped in the same *nn* individuals. Denote *G*_1_ as the *n* ×*p*_1_ genotype matrix for *p*_1_ variants in the first gene region, and *G*_2_ as the *n* × *p*_2_ genotype matrix for *p*_2_ variants in the second gene region.

### Multi-phenotype and G×G interaction unit-based test

Before presenting the proposed test for associations between multiple phenotypes and G×G interactions, we define the interaction term. Let *S* be the *n* × *p*_*C*_ interaction matrix, where *p*_*C*_ = *p*_1_ × *p*_2_ representing pairwise interactions between variants from two gene regions (*p*_1_ and *p*_2_ denote the numbers of variants in each region). For convenience, denote 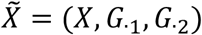. We model the association between the *k*-th phenotype and 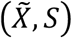 with a generalized linear model:

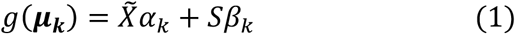

where ***μ***_***k***_ = (*μ*_1*k*_, …, *μ*_*nk*_)^*T*^ denotes the estimated mean of ***y***_***k***_ = (*y*_1*k*_, *y*_2*k*_, …, *y*_*nk*_)^*T*^ under the null hypothesis. For *K* phenotypes, the generalized linear model can be extended to

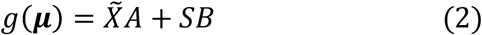

where *A* = (*α*_1_, …, *α*_*k*_) is the coefficient matrix for 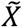 and *S* = (*β*_1_, …, *β*_*k*_) is the *p*_*C*_ × *K* coefficient matrix for the interaction term *S*. Our goal is to test the null hypothesis *H*_0_: *S* = 0, *i*.*e*., no association between G×G interaction terms and the phenotypes. To this end, we construct the following score test statistic:

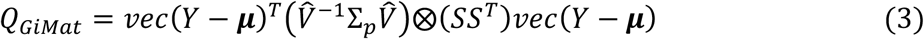

where ***μ*** = (***μ***_**1**_, …, ***μ***_***n***_)^*T*^, ⨂ denotes the Kronecker product, *vec* (·) is the vector opterator that stacks the columns of a matrix into a vector, and 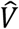 denotes the residual covariance of *Y* under *H*_0_. Under *H*_0_, *Q*_*GiMat*_ asymptotically follows a weighted sum of chi-square distributions with one degree of freedom: 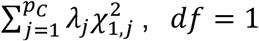. Specifically, λ (*j* = 1, …, *p*) are eigenvalues of 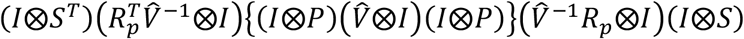 where 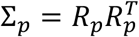 and *P* is the block diagonal matrix, *diag* (*P*_1_, …, *P*_*k*_) with each block matrix 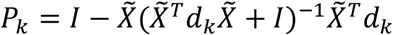, where *d*_*k*_ = *diag* (*μ*_*k*_(1 − *μ*_*k*_)). The *p*-values can be estimated using Davies’ method^44^.

The matrix Σ_*p*_ specifies the relationship among the effect sizes of genetic interactions on each of the phenotypes, and can be any semi-definite matrix. In GiMat, we extend the choices of Σ_*p*_ from Multi-SKAT^35^ to accommodate multiple dichotomous phenotypes and G×G interactions.

### Homogeneous kernel (*Hom*)

The *Hom* kernel modelling the phenotype covariance matrix Σ_*p*_ assumes that the effect sizes of G × G interactions act in the same direction across all phenotypes. Under this assumption, Σ_*p,Hom*_ = Γ_*K*_ Γ _*K*_^*T*^, where Γ_*K*_ = (1, …, 1)^*T*^.

## Heterogeneous kernel (*Het*)

In contrast, the *Het* kernel allows the effect sizes of G×G interactions to be uncorrelated or even act in opposite directions across phenotypes. Specifically, Σ_*p*_ is constructed as Σ_*p,Het*_ = *K*, where *K* = *diag*(1, …, 1).

### Phenotype covariance kernel (*PhC*)

Alternatively, Σ_*p*_ can be specified proportional to the estimated residual covariance across all phenotypes. It can be expressed as 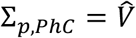, where 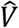 denotes the estimated covariance matrix among phenotypes. This kernel assumes that the covariance of genetic interaction effects is proportional to the residual phenotypic covariance after adjusting for non-genetic covariates.

## Gene association with multi-trait test (*GAM*)

We also include 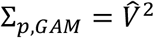 which has been proposed as an alternative kernel^35^ and is mathematically equivalent to the phenotype kernels used in GAMuT^13^ and MSKAT^15^.

### Minimum *p*-values omnibus test (*minP*)

Choosing Σ_*p*_ solely from prior biological knowledge risks power loss if the assumed Σ_*p*_ structure is inconsistent with the true effect pattern^16,45^. To improve robustness, GiMat implements an omnibus test by taking the minimum *p*-value across all kernel-specific tests. While permutation or perturbation methods can be applied to calculate the Monte‐Carlo *p*-values^14,46^, they are computationally intensive. To overcome this limitation, we adopt a fast Copula-based *p*-value calculation strategy^35^, and it requires fewer resampling steps to accurately estimate *p*-values.

Specifically, let *p*_*h*_ represents the *p*-values for *Q*_*GiMat*_ with given *h*-th Σ_*p*_, *h* = 1, …, *H*, and *T*_*p*_ = (*p*_1_, …, *p*_*H*_)^*T*^ denotes the *H* × 1 vector of *p*-values from a number of *H* GiMat tests. Assuming that the joint distribution of *T*_*p*_ denotes multivariate *t*-distribution with an estimated correlation structure, a *t*-Copula is used to approximate their joint multivariate distribution of *T*_*p*_ by modelling the marginal distribution of each statistic *Q*_*GiMat*_ with different Σ_*p*_ and respective correlation structure^47,48^. Consequently, the omnibus *p*-value, *GiMat* _*minP*_, can be efficiently computed from the distribution of the assumed *tt*-Copula, providing an accurate and efficient alternative to Monte‐Carlo resampling.

### Simulation design

We conducted simulation studies to evaluate GiMat’s performance in terms of empirical type I error control and statistical power. Haplotype data from the 1000 Genomes Project^49^ served as the reference panel. Using HapGen2^50^, we simulated 30,000 haplotypes spanning a 5Mb region on chromosome 18 and focused on interactions between common variants (MAF > 1%) for two dichotomous phenotypes. GiMat tests were conducted with five kernel specifications: *Q*_*GiMat,Het*_, *Q*_*GiMat,Hom*_, *Q*_*GiMat,Phc*_, *Q*_*GiMat,GAM*_, and *Q*_*GiMat,minP*_, and their respective type I error rates and power were assessed.

### Type I error evaluation

To assess type I error control under various scenarios, we simulated 10,000 independent genotype datasets, each comprising 5,000 individuals. For each dataset, two continuous phenotypes were first generated under the model:

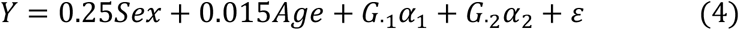

where *Y* = (*y*_1_, *y*_2_) and ε denotes a vector of residual errors drawn from a multivariate normal distribution *MVN* (0, Σ) with mean zero and covariance matrix Σ that controls the correlation structure between phenotypes. To evaluate robustness, we examined three correlation scenarios by setting the correlation between *y*_1_ and *y*_2_ to 0.3, 0.5, or 0.75.

Two non-genetic covariates were included: *Age* was a continuous variable sampled from a normal distribution with mean 48 and standard deviation 3. *Sex* was a binary variable sampled from a Bernoulli distribution with a probability of 50% for male.

To enhance the generalizability of our simulations, we considered two genetic-effect scenarios: (a) without main genetic effects (*α*_1_ = *α*_2_ = 0) and (b) with main genetic effects, where *G*_1_ was associated with phenotype *y*_1_ and *G*_2_ with phenotype *y*_2_. Specifically, one SNP was assigned to *G*_1_ and two SNPs to *G*_2_.

Dichotomous phenotypes were then derived by thresholding the continuous phenotypes to achieve specified disease prevalences of 0.05, 0.10, 0.20, and 0.50. Empirical type I error rates were estimated as the proportion of *p*-values ≤ 0.05, 0.01, 0.005, 0.001, and 0.0005 across simulations.

### Statistical power

For power evaluation, we generated two continuous phenotypes under two genetic scenarios: (i) without main genetic effects and (ii) with main genetic effects. Using the simulated genotype data, phenotypes were generated according to:

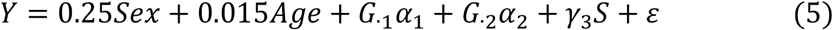

where *Y* = (***y***_**1**_, ***y***_**2**_) and ε denotes the error term following a multivariate normal distribution *MVN*(0, Σ) with mean zero and correlation matrix Σ. It is noted that the correlation between phenotypes was set to 0.5. *S* = (*S*_1_, …, *S*_*k*_), representing the G×G interaction term.

In both genetic scenarios, a subset of G×G interaction terms were assumed to influence the phenotypes. We varied (a) the proportion of causal interactions and (b) the direction of their effects. Specifically, (a) 20%, 60%, and 100% of G×G interactions were designated as causal variants associated with both phenotypes; (b) two effect configurations were considered: first, all causal variants exert positive effects, and second, half exert positive and half exert negative effects. Effect sizes for causal G×G interactions were fixed at γ_3_ = 0.2. Main genetic effects for causal variants, *α*_*j*_ (*j* = 1,2), followed a 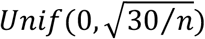 distribution^14^.

To contrast the *Hom* and *Het* kernels, phenotypes were also simulated under the mixed-direction setting. For example, with 20% of G × G interactions designated as causal, the first 10% exerted positive effects on the first phenotype and negative effects on the second phenotype, while the remaining 10% exhibited the opposite pattern. Dichotomous phenotypes were then derived using the same thresholding approach as in the type I error evaluation, fixing disease prevalence at 20%.

For each simulation configuration, we conducted 1,000 replicates. Statistical power was defined as the proportion of replicates with *p*-values below the nominal significance level.

## Detection of G×G Interactions in T2D-HTN Comorbidity Using UK Biobank Data

### Data preprocessing

We utilized genotype and phenotype data from the UK Biobank to evaluate GiMat’s ability to detect G×G interactions underlying the comorbidity of T2D and HTN. The UK Biobank is a nation-scale biomedical database containing genetic, lifestyle, and health information on approximately half a million participants^51^.

First, we identified genomic variants associated with T2D or HTN from publicly available GWAS results (http://www.nealelab.is/uk-biobank/) and the GWAS Catalog^52^, restricting analyses to individuals of white British ancestry. We retained variants with reported *p*-values<5×10^−8^ and MAF > 5%.

Second, to map phenotype-associated SNPs to their cis-regulated genes, we used genome-wide fine-mapping results from Gazal et al.^53^, which cover 19,476,620 SNPs and their linked genes in the UK Biobank (MAF ≥ 0.5%). We kept SNP-gene associations with a cS2G linking score ≥ 0.5.

Third, we prioritized potentially deleterious SNPs using Ensembl Variant Effect Predictor (VEP)^54^. For each SNP, we obtained three predictive scores: Sorting Intolerant From Tolerant (SIFT), Polymorphism Phenotyping (PolyPhen), and Combined Annotation Dependent Depletion (CADD). These scores can quantify the potential impact of each SNP on gene function. SNPs that failed to meet thresholds for any of these predictors were excluded from further analysis.

Finally, we extracted the array-genotype data of the selected SNPs from the UK Biobank. SNPs linked to each gene were organized into separate text files, and associated phenotypes were annotated based on the linking SNPs, classifying genes as associated with T2D, HTN, or both.

### Sample Stratification

We analyzed individual-level data from 337,205 white British participants in the UK Biobank, incorporating both participant characteristics and SNP-array genotypes to test associations between G×G interactions and phenotypes. Participants were categorized into four groups: (1) both T2D and HTN, (2) T2D only, (3) HTN only, and (4) neither disease. To investigate whether G × G interactions influencing disease pathogenesis vary by age of disease onset, we stratified participants with both T2D and HTN into four subgroups defined by the difference between age at T2D diagnosis and age at HTN diagnosis: (−∞, −5], (−5, 0], (0, 5], and (5, +∞) years. For each subgroup, cases (participants with both diseases) were matched to controls drawn from the remaining three phenotype groups on sex, age at diagnosis, and age at recruitment. When multiple controls met these criteria, we selected the individual with the smallest Euclidean distance between the top two principal components (PCs) derived from genome-wide principal component analysis. The distribution of participants across subgroups is provided in Supplementary Table 3. GiMat was then applied independently within each subgroup to identify G×G interactions potentially involved in T2D-HTN comorbidity.

Quantile-quantile (Q-Q) plots of these resulting *p*-values (Supplementary Fig. 3) showed most points aligning closely with the expected null distribution, with only slight inflation in the tails. We quantified this inflation using the genomic inflation factor *λ*_*IF*_ for each kernel across the four age-difference subgroups. In the (0, 5] subgroup, the *Het* test exhibited moderate inflation (*λ*_*IF*_ = 1.295) but was slightly deflated in the other subgroups (*λ*_*IF*_ = 0.984, 0.992, and 0.868). The *minP* test showed deflation across all subgroups. Both the *Hom* and *PhC* kernels exhibited inflation in the (0, 5] subgroup (*λ*_*IF*_ = 1.307 and 1.149) but were deflated elsewhere. The *GAM* test displayed deflation only in the (5, +∞) subgroup (*λ*_*IF*_ = 0.813).

To identify which age-difference subgroup was most informative for detecting significant G×G interactions, we compared the distributions of significant *p*-values obtained from the same kernel tests across subgroups. For each subgroup, we first selected G × G interactions that GiMat tested to be significantly associated with T2D and HTN in each subgroup and transformed their *p*-values to –log_10_(*p*-values). We then matched –log_10_(*p*-values) between any two subgroups based on G×G interaction names and the tested kernel. We then applied the Wilcoxon signed-rank test to determine whether the distributions of –log_10_(*p*-values) differed significantly between paired subgroups (Supplementary Table 5). This test yields both a *p*-value and an effect size. A *p*-value < 0.05 indicates that a significant difference in distributions. The effect size reflects the direction and magnitude of the difference: a positive value indicates that interactions in the first subgroup tend to have smaller (more significant) *p*-values than those in the second subgroup, whereas a negative value indicates the reverse.

### Validation of Significant G×G Interactions through Co-expression Analysis

To examine whether the significant G×G interactions identified in Table 1 reflect coordinated transcriptional activity, we analyzed RNA-seq data from 54 non-diseased tissues across nearly 1,000 individuals from the Genotype-Tissue Expression (GTEx) project^23^. Data for this analysis were downloaded from the GTEx Portal^23^ on 17 July 2024. For each G × G interaction, the Pearson Correlation Coefficient (PCC) of gene expressions was calculated separately within each tissue (Supplementary Fig. 2). Statistical significance of each PCC was evaluated using hypothesis testing with false discovery rate (FDR) correction to account for multiple comparisons.

## Data Availability

All data produced in the present study are available upon reasonable request to the authors.

## Data availability

The phenotypic and genetic data from the UK Biobank used in this study are under the application license 78814. Researchers can apply for access at https://www.ukbiobank.ac.uk/enable-your-research/apply-for-access. Further details on the UK Biobank and its datasets can be found at https://www.ukbiobank.ac.uk/.

## Code availability

The GiMat framework is freely available as an R library at https://github.com/SiruRooney/GiMat.

## Acknowledgements

This work was supported by the National Natural Science Foundation of China (No. 32470720), Basic Research Center, Innovation Program of Chinese Academy of Agricultural Sciences (No. CAAS-BRC-FNH-2025-02), and the Agricultural Science and Technology Innovation Program (ASTIP) awarded to Gengjie Jia. Support to Yu Li was provided by the Chinese University of Hong Kong (CUHK, No. 4937025, 4937026, 5501517, 5501329, 8601603, 8601663, and SHIAE BME-p1-24), Research Grants Council of the Hong Kong Special Administrative Region, China (Hong Kong SAR, No. CUHK 24204023), Innovation and Technology Commission of the Hong Kong SAR (No. GHP/065/21SZ), and CUHK Ideaboost (No. IDBF23ENG05). The funders had no involvement in the study design, data collection or analysis, decision to publish, or manuscript preparation.

## Author contributions

S.W. developed the GiMat method and software; S.W. and X.Z. performed statistical analyses. S.W., X.Z., G.J., and Y.L. wrote and revised the manuscript; G.J. conceived and designed the project. All authors read and approved the final version of the manuscript.

