## Supplementary material for "A statistical framework to identify gene-gene interactions underlying multiple dichotomous phenotypes from genotype data"

**Supplementary Table 1. Empirical type I error ratios for testing associations between moderately correlated phenotypes and G×G interactions, without (top panel) and with (bottom panel) main genetic effects.** Each cell reports the ratio of the empirical type I error to the nominal significance level $\theta$. Type I error was estimated as the proportion of *p*-values < 0.05, 0.01, 0.005, 0.001, and 0.0005 across 10,000 simulation replicates at disease prevalence levels of 5%, 10%, 20%, and 50%.

| *θ* | *Het* | *Hom* | *PhC* | *GAM* | *minP* |
|---|---|---|---|---|---|
| Without main genetic effects | | | | | |
| Prevalence 5% | | | | | |
| 0.05 | 0.354 | 0.358 | 0.352 | 0.350 | 0.246 |
| 0.01 | 0.360 | 0.310 | 0.330 | 0.350 | 0.260 |
| 0.005 | 0.420 | 0.340 | 0.420 | 0.460 | 0.260 |
| 0.001 | 0.400 | 0.300 | 0.400 | 0.500 | 0.400 |
| 0.0005 | 0.800 | 0.600 | 0.600 | 0.800 | 0.800 |
| Prevalence 10% | | | | | |
| 0.05 | 0.466 | 0.500 | 0.480 | 0.466 | 0.288 |
| 0.01 | 0.420 | 0.410 | 0.410 | 0.410 | 0.230 |
| 0.005 | 0.360 | 0.320 | 0.340 | 0.320 | 0.280 |
| 0.001 | 0.600 | 0.700 | 0.700 | 0.700 | 0.500 |
| 0.0005 | 1.000 | 0.600 | 1.000 | 1.000 | 1.000 |
| Prevalence 20% | | | | | |
| 0.05 | 0.612 | 0.642 | 0.622 | 0.636 | 0.362 |
| 0.01 | 0.450 | 0.510 | 0.490 | 0.470 | 0.260 |
| 0.005 | 0.420 | 0.380 | 0.420 | 0.400 | 0.260 |
| 0.001 | 0.300 | 0.300 | 0.300 | 0.300 | 0.300 |
| 0.0005 | 0.600 | 0.600 | 0.600 | 0.600 | 0.600 |
| Prevalence 50% | | | | | |
| 0.05 | 0.720 | 0.744 | 0.722 | 0.724 | 0.460 |
| 0.01 | 0.740 | 0.830 | 0.770 | 0.760 | 0.43 |
| 0.005 | 0.680 | 0.760 | 0.740 | 0.680 | 0.480 |
| 0.001 | 1.300 | 1.100 | 1.200 | 1.200 | 0.600 |
| 0.0005 | 0.600 | 0.800 | 0.600 | 0.600 | 0.600 |
| With main genetic effects | | | | | |
| Prevalence 5% | | | | | |
| 0.05 | 0.014 | 0.014 | 0.014 | 0.014 | 0.012 |
| 0.01 | 0.030 | 0.030 | 0.030 | 0.030 | 0.010 |
| 0.005 | 0.020 | 0 | 0.020 | 0.020 | 0.020 |
| 0.001 | 0 | 0 | 0 | 0 | 0 |
| 0.0005 | 0 | 0 | 0 | 0 | 0 |
| Prevalence 10% | | | | | |
| 0.05 | 0.016 | 0.020 | 0.020 | 0.018 | 0.014 |

| | | | | | |
|---|---|---|---|---|---|
| 0.01 | 0.030 | 0.010 | 0.010 | 0.03 | 0.01 |
| 0.005 | 0.020 | 0.020 | 0.020 | 0.020 | 0 |
| 0.001 | 0 | 0 | 0 | 0 | 0 |
| 0.0005 | 0 | 0 | 0 | 0 | 0 |
| | | | Prevalence 20% | | |
| 0.05 | 0.040 | 0.048 | 0.046 | 0.042 | 0.024 |
| 0.01 | 0.040 | 0.050 | 0.050 | 0.050 | 0.030 |
| 0.005 | 0.020 | 0.040 | 0.040 | 0.020 | 0 |
| 0.001 | 0 | 0 | 0 | 0 | 0 |
| 0.0005 | 0 | 0 | 0 | 0 | 0 |
| | | | Prevalence 50% | | |
| 0.05 | 0.096 | 0.112 | 0.102 | 0.094 | 0.060 |
| 0.01 | 0.050 | 0.040 | 0.060& | 0.060 | 0.030 |
| 0.005 | 0.060 | 0.040 | 0.040 | 0.060 | 0.040 |
| 0.001 | 0.100 | 0.100 | 0.100 | 0.100 | 0 |
| 0.0005 | 0 | 0 | 0 | 0 | 0 |

**Supplementary Table 2. Empirical type I error rates for testing associations between G × G interactions and multiple dichotomous phenotypes under different phenotype correlation patterns.** Each cell shows the ratio of the empirical type I error rate to the significance level $\theta$. Type I error was estimated as the proportion of *p*-values < 0.05, 0.01, 0.005, 0.001, and 0.0005 based on 10,000 simulation replicates. Top panel: low-correlated phenotypes; Bottom panel: high-correlated phenotypes.

| With the main genetic effects and at the disease prevalence of 20% | | | | | |
|---|---|---|---|---|---|
| *θ* | *Hom* | *Het* | *PhC* | *GAM* | *minP* |
| Under the low-correlated phenotypes | | | | | |
| 0.05 | 0.062 | 0.066 | 0.068 | 0.064 | 0.048 |
| 0.01 | 0.060 | 0.050 | 0.070 | 0.060 | 0.030 |
| 0.005 | 0.060 | 0.060 | 0.060 | 0.060 | 0.040 |
| 0.001 | 0.100 | 0.100 | 0.100 | 0.100 | 0.100 |
| 0.0005 | 0.200 | 0.200 | 0.200 | 0.200 | 0.200 |
| Under the high-correlated phenotypes | | | | | |
| 0.05 | 0.028 | 0.034 | 0.026 | 0.026 | 0.022 |
| 0.01 | 0.040 | 0.040 | 0.040 | 0.040 | 0.020 |
| 0.005 | 0.040 | 0.040 | 0.040 | 0.040 | 0.020 |
| 0.001 | 0 | 0 | 0 | 0 | 0 |
| 0.0005 | 0 | 0 | 0 | 0 | 0 |

**Supplementary Table 3. Participant counts by age-difference subgroup.** Each subgroup is defined by an integer interval representing the difference between age at T2D onset and age at HTN onset for individuals within that subgroup.

| Age-difference subgroup | Participant count |
|---|---|
| $(-\infty, -5]$ | 2,718 |
| $(-5, 0]$ | 6,310 |
| $(0, 5]$ | 4,351 |
| $(5, +\infty)$ | 5,379 |

**Supplementary Table 4. Significant G×G interactions that overlap with molecular interactions reported in IntAct and STRING databases**. An asterisk (*) denotes *p*-value < 0.05, indicating that the corresponding G×G interaction was significantly associated with both T2D and HTN by GiMat. Each row lists a G×G interaction, where the first gene in the “G×G Interaction (SNPs)” column is associated with T2D and the second with HTN. SNP loci from which array data were derived are shown in brackets after each gene. **Age-difference group** indicates the participant subgroup in which GiMat identified the significant *p*-value, defined by an integer interval representing the difference between T2D and HTN onset ages. **Interaction Type (DB)** specifies the type of molecular interaction between G×G interactions. IntAct provides detailed interaction types and an MIscore reflecting the confidence level of the reported interaction. STRING indicates the presence of an association without additional interaction details. According to IntAct definitions, "direct interaction" refers to molecules in direct contact and "physical association" denotes molecules within the same physical complex. The databases providing supporting evidence are noted in brackets. **Kernel abbreviations**: *Het*, heterogeneous kernel; *Hom*, homogeneous kernel; *PhC*, phenotype-covariance kernel; *GAM*, gene association with multi-trait test; *minP*, minimum *p*-values omnibus test.

| G×G Interaction (SNPs) | Age-difference group | Interaction Type (DB) | *Het* | *Hom* | *PhC* | *GAM* | *minP* |
|---|---|---|---|---|---|---|---|
| APOE (rs429358)<br>MTHFR (rs1801131, rs2066462) | (5, +∞) | None (STRING) | $4.56\times10^{-3*}$ | $3.67\times10^{-3*}$ | $1.17\times10^{-2*}$ | $3.71\times10^{-2*}$ | $7.05\times10^{-3*}$ |
| INS (rs3842752)<br>HLA-DQB1 (rs1130399, rs1130398, rs9274407) | (-5, 0] | None (STRING) | $2.38\times10^{-2*}$ | $2.94\times10^{-2*}$ | $2.12\times10^{-2*}$ | $1.84\times10^{-2*}$ | $3.97\times10^{-2*}$ |
| HLA-DQA1 (rs13266634)<br>HLA-DQB1 (rs1130399, rs1130398, rs9274407) | (0, 5] | Direct interaction (STRING/ IntAct, MIscore=0.82) | $1.97\times10^{-2*}$ | $3.13\times10^{-2*}$ | $1.67\times10^{-2*}$ | $2.41\times10^{-2*}$ | $3.26\times10^{-2*}$ |
| INS (rs3842752)<br>BAG6 (rs4354268, rs74990397) | (0, 5] | Physical association (IntAct, MIscore=0.56) | $4.49\times10^{-2*}$ | $3.79\times10^{-2*}$ | $9.04\times10^{-2}$ | $4.07\times10^{-2*}$ | $7.23\times10^{-2}$ |

**Supplementary Table 5. Comparison of *p*-value distributions for significant G×G interactions across age-difference subgroups using the Wilcoxon signed-rank test.** This table summarizes pairwise comparisons of significant G×G interaction signals detected by GiMat across age-difference subgroups. For each subgroup, G×G interactions significantly associated with T2D and HTN were selected, and their *p*-values were transformed to $-\log_{10}$(*p*-values). For each pair of subgroups, $-\log_{10}$(*p*-values) were matched by identical gene interaction names and kernel type. The Wilcoxon signed-rank test was then applied to determine whether the distributions of $-\log_{10}$(*p*-values) differed significantly between subgroups. **Age-difference group** lists the paired age-difference subgroups used for comparison. **Kernel type** indicates the phenotype kernel of GiMat used for the corresponding *p*-value subgroup. ***p*-values of difference** gives the *p*-values obtained from the Wilcoxon signed-rank test, assessing whether the distributions of significant $-\log_{10}$(*p*-values) differ between subgroups; asterisks (*) indicate statistical significance at the 0.05 level. **Effect size** reports the overall shift in $-\log_{10}$(*p*-value) distributions between paired subgroups: positive values indicate stronger significance (smaller *p*-values) in the first subgroup relative to the second, and negative values indicate the reverse.

| Age-difference group | Kernel type | *p*-value of difference | Effect size |
|---|---|---|---|
| (-∞, -5] & (-5, 0] | *Het* | 0.807 | 0.028 |
| | *Hom* | 0.613 | 0.055 |
| | *PhC* | 0.629 | 0.049 |
| | *GAM* | 0.723 | 0.044 |
| | *minP* | 0.974 | 0.002 |
| (-∞, -5] & (0, 5] | *Het* | 0.046* | -0.179 |
| | *Hom* | 0.045* | -0.204 |
| | *PhC* | 0.096 | -0.145 |
| | *GAM* | 0.068 | -0.184 |
| | *minP* | 0.104 | -0.127 |
| (-∞, -5] & (5, +∞) | *Het* | 0.462 | 0.058 |
| | *Hom* | 0.237 | 0.116 |
| | *PhC* | 0.776 | -0.026 |
| | *GAM* | 0.315 | 0.090 |
| | *minP* | 0.486 | 0.052 |
| (-5, 0] & (0, 5] | *Het* | 0.008* | -0.235 |
| | *Hom* | 0.005* | -0.281 |
| | *PhC* | 0.027* | -0.217 |
| | *GAM* | 0.007* | -0.263 |
| | *minP* | 0.032* | -0.184 |
| (-5, 0] & (5, +∞) | *Het* | 0.666 | -0.041 |
| | *Hom* | 0.551 | -0.059 |
| | *PhC* | 0.602 | -0.049 |
| | *GAM* | 0.632 | -0.049 |
| | *minP* | 0.832 | -0.017 |
| (0, 5] & (5, +∞) | *Het* | 0.015* | 0.208 |
| | *Hom* | 0.008* | 0.247 |
| | *PhC* | 0.100 | 0.142 |
| | *GAM* | 0.015* | 0.229 |
| | *minP* | 0.041* | 0.152 |

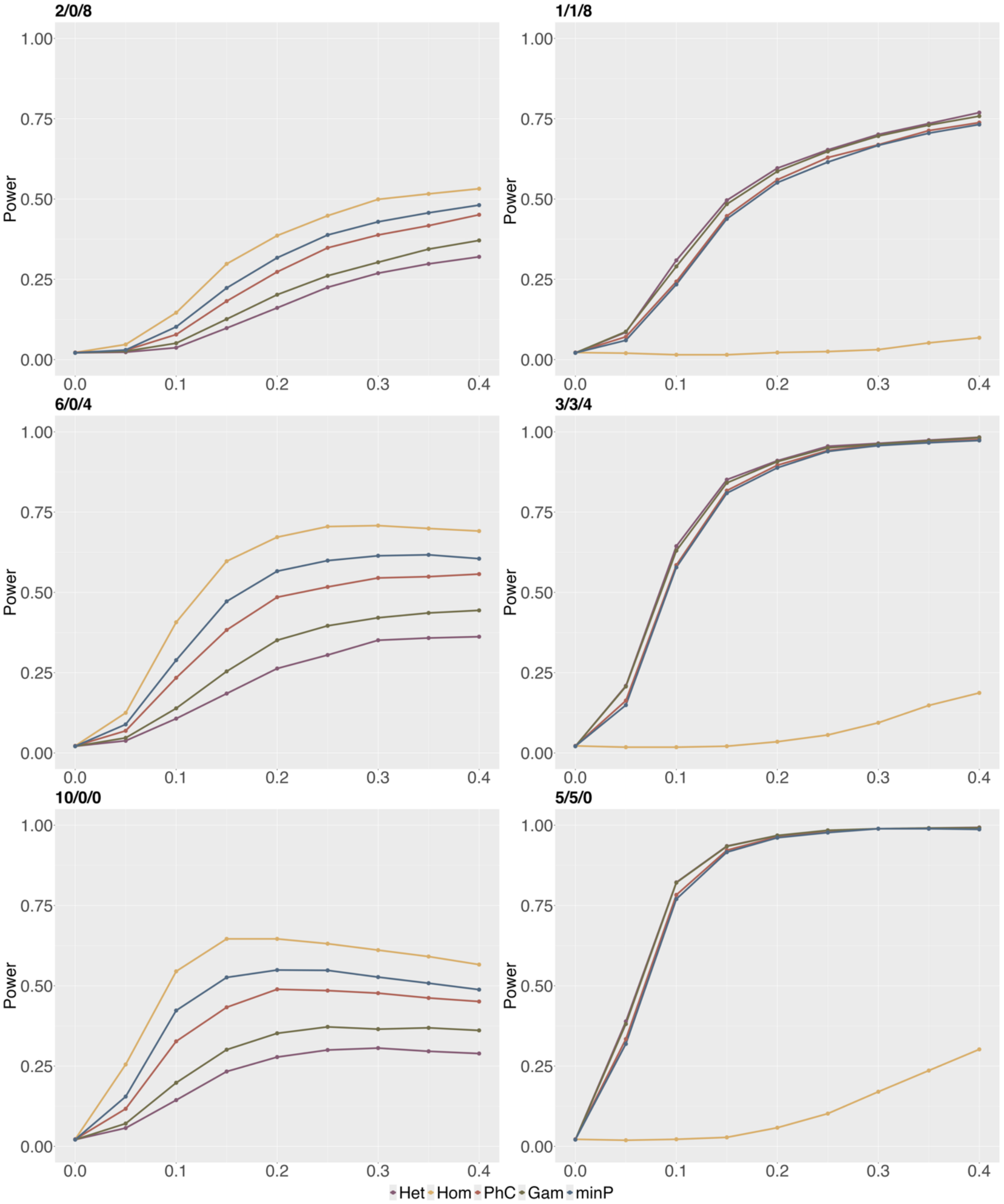


**Supplementary Fig. 1. Empirical power curves for detecting G×G interactions (at the $\theta = 0.025$ significance level) in scenarios where both interaction effects and main genetic effects contribute to distinct phenotypes**. The format of “+/-/0” indicates interaction variants with positive, negative, and null effects. The left panel shows the empirical power curves for scenarios with 2/0/8, 6/0/4 and 10/0/0 causal variants, while the right panel shows the empirical power curves for scenarios with 1/1/8, 3/3/4 and 5/5/0 causal variants. The X-axis represents the effect sizes of causal interactions ranging from 0 to 0.4 The y-axis shows statistical power, defined as the proportion of the 1,000 replicates with *p*-values below the nominal significance level.

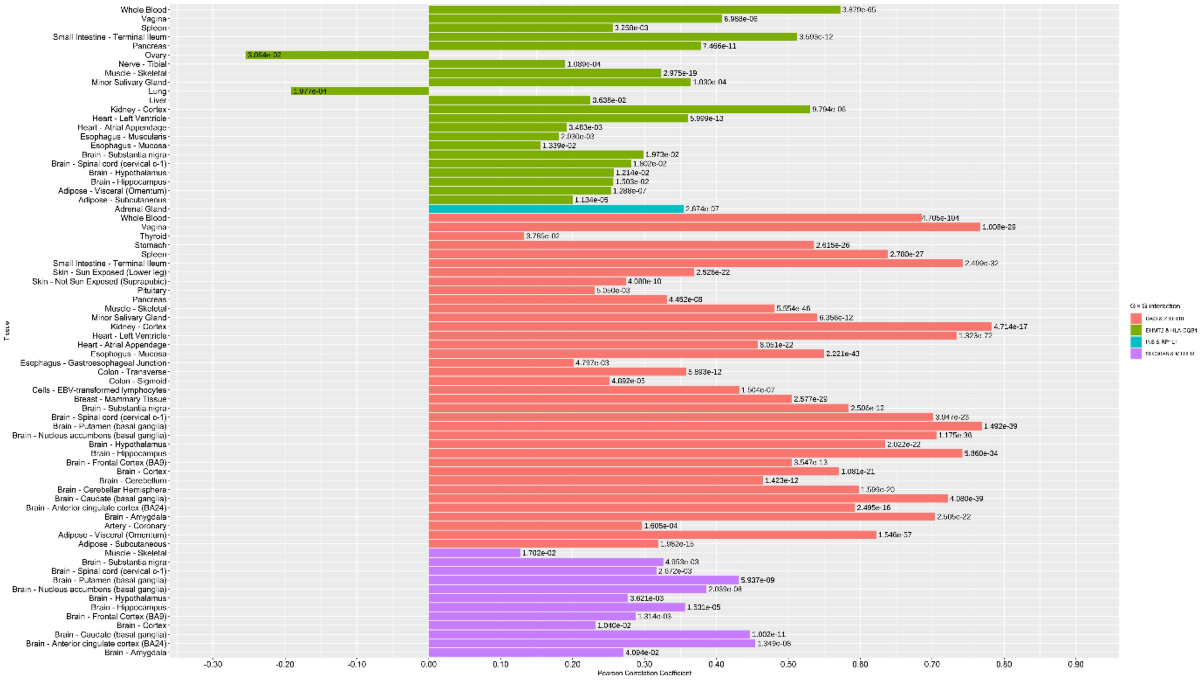

**Supplementary Fig. 2. Tissues in which four G×G interactions exhibit significant co-expression relationships**. The bar length along the x-axis indicates the Pearson Correlation Coefficient (PCC) for each gene pair within a specific tissue. The corresponding *p*-value displayed on the right of each bar has been adjusted for false discovery rate (FDR), reflecting the statistical significance of the PCCs. FDR correction is applied to control for false positives due to multiple testing.

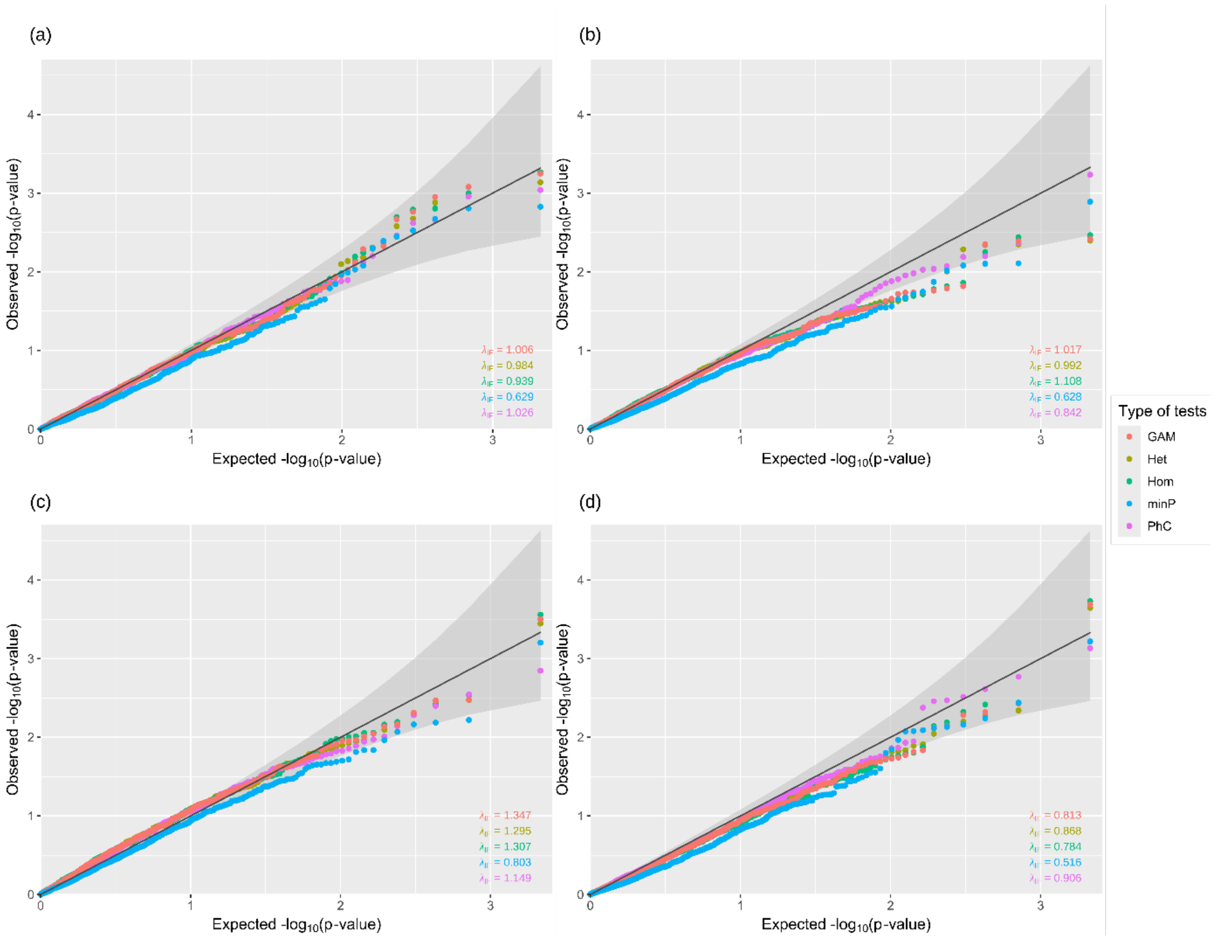


**Supplementary Fig. 3. Quantile-quantile (Q-Q) plots of *p*-values derived from the *Het*, *Hom*, *PhC*, *GAM*, and *minP* tests, using data from four age-difference subgroups.** Panels (a), (b), (c), and (d) show the *p*-values from these tests for detecting G×G interactions within the subgroups corresponding to age-difference intervals of (-∞, -5], (-5, 0], (0, 5], and (5, +∞), respectively. The lower right corner of each subplot displays the inflation factor $\lambda_{IF}$, which quantifies the extent of false positive inflation in statistical tests. It is calculated as the ratio of the observed test statistics to their expected values under the null hypothesis. Abbreviations: *Het*, heterogeneous kernel; *Hom*, homogeneous kernel; *PhC*, phenotype covariance kernel; *GAM*, gene association with multi-trait test; *minP*, minimum *p*-values omnibus test.
